# Photoreceptor outer segment reflectivity with ultrahigh resolution visible light optical coherence tomography in systemic hydroxychloroquine use

**DOI:** 10.1101/2024.09.10.24313265

**Authors:** Anupam K. Garg, Jingyu Wang, Bailee Alonzo, Ji Yi, Amir H. Kashani

**Affiliations:** Wilmer Eye Institute, Johns Hopkins University, Baltimore, MD, USA; Department of Biomedical Engineering, Johns Hopkins University, Baltimore, MD, USA; Department of Biomedical Engineering, Johns Hopkins Hospital, Baltimore, MD, USA

## Abstract

**Purpose:** To evaluate outer retinal organization in normal subjects and those using hydroxychloroquine (HCQ) with ultrahigh resolution visible light optical coherence tomography (VIS-OCT).

**Methods:** Forty eyes of 22 adult subjects were recruited from a tertiary care retina practice including controls (20 eyes, 12 subjects, mean age 40±22yrs, mean logMAR BCVA 0.19, 90% female) and subjects with a history of HCQ use (20 eyes, 10 subjects, mean age 62±17yrs, mean logMAR BCVA 0.03, 67% female). Each subject was imaged using a custom-built VIS-OCT device (axial resolution 1.3μm) and FDA-approved OCT devices.

**Results:** Using VIS-OCT, control subjects demonstrate 5 and 6 hyperreflective bands in the foveal and parafoveal regions, respectively, between the outer nuclear layer and Bruch’s membrane. These bands demonstrate intensity profiles complementary to the known histopathologic distribution of rods and cones. In comparison to controls, subjects taking HCQ demonstrate blunting of all bands, particularly bands 2-4. In all cases of suspected or known toxicity, VIS-OCT demonstrated attenuation of band 3i and in no cases was there attenuation of other bands that was more severe than band 3i, suggesting that changes in the reflectivity of Band 3i may be the earliest identifiable sign of HCQ toxicity.

**Conclusions:** VIS-OCT of the outer retina demonstrates a unique outer retinal banding pattern corresponding to photoreceptor density profiles. There is a notable attenuation of the photoreceptor outer segment reflectivity profile associated with early HCQ toxicity. This finding may be an early, and possibly reversible, sign of HCQ toxicity, primarily impacting the cones.

## Introduction

Hydroxychloroquine (HCQ) is a well-described anti-inflammatory and anti-malarial medication with relatively rare but devastating toxic effects on the retina^1^. Although a benign drug in general, the potential for this serious side-effect causes significant problems in the management of patients who are on the medication. The exact pathophysiology of the side-effect is not well understood however several lines of evidence suggest that toxicity at the level of the retinal pigment epithelium and, secondarily the photoreceptor, is likely. *In vitro* studies have demonstrated that HCQ inhibits protein synthesis^2^ and lysosomal function^3^ in RPE. Toxicity manifests in subjects with prolonged HCQ exposure as thinning of the outer retina and loss of the retinal pigment epithelium in a bulls-eye pattern^4^. However, even subjects without any clinical findings or subjective symptoms can demonstrate subclinical signs of toxicity such as modest retinal thinning on OCT and decreased signal amplitude on mfERG^5,6^. Careful analysis of OCT images has suggested these changes are caused by loss of the ellipsoid zone^7^ and/or changes in the photoreceptor outer segments^8^. While severe changes may be seen with retinal thickness maps^9^, the resolution of commercially available OCT devices is not sufficient to easily and reliably assess subclinical changes in the outer segment banding patterns on single OCT volumes^10^. To overcome this limitation, serial OCT imaging has been used to detect very subtle paracentral thinning patterns that seem strongly correlated with toxicity^6^. This finding suggests that higher resolution imaging of the photoreceptor/RPE complex may reveal even earlier signs of toxicity.

While most commercial devices utilize near-IR wavelength light, ultrahigh resolution visible light OCT (VIS-OCT) utilizes shorter wavelengths of light to enable higher axial resolution (∼1 µm) compared to conventional OCT devices^11^. Recent technological advances have enabled the development of VIS-OCT devices for human retinal imaging, enabling characterization of both inner and outer retinal layers in greater detail than previously possible in human subjects^12–15^ and animal models^16,17^. Prior work has demonstrated that VIS-OCT reveals outer retinal banding morphology that was not readily visible with conventional devices in control subjects without a known history of retinal pathology, including Bruch’s membrane and sub-bands in photoreceptor outer segments^18^. Similar banding morphology has also been recently demonstrated using ultrahigh resolution SD-OCT prototype devices^19,20^. *In vivo* characterization of these fine details and their changes in retinal disease may provide valuable insights into disease pathophysiology and management. Given that the outer retina is primarily affected in early HCQ toxicity, VIS-OCT is a promising imaging modality to enable early detection of HCQ toxicity. In this study we used a custom-built VIS-OCT device to identify subtle outer retinal changes in HCQ toxicity not visible with conventional OCT devices.

## Methods

### Recruitment of Subjects

Both eyes of adult subjects with a history of hydroxychloroquine use were prospectively recruited from a tertiary care retina practice for retinal imaging with a custom-built dual-channel VIS-OCT device with an axial resolution of 1.3μm (see Wang et al., 2022 for technical details^18,21^) (**Figure 1**). Each subject was also imaged using a commercial swept-source OCT imaging device. The most recent hepatic and renal function testing results of each subject were also obtained. Control subjects were recruited from the retina clinics if they had no vision threatening retinal or ocular disease in at least one eye. Information regarding subjects is summarized in **Table 1**. All subjects provided informed consent according to a human subject protocol approved by the Johns Hopkins Medicine Institutional Review Boards (IRB) and in accordance with the principles of the Declaration of Helsinki. Subjects with significant media opacity, poor signal quality, or inability to fixate sufficiently to obtain at least one high quality foveal line scan were excluded.

**Figure 1:**
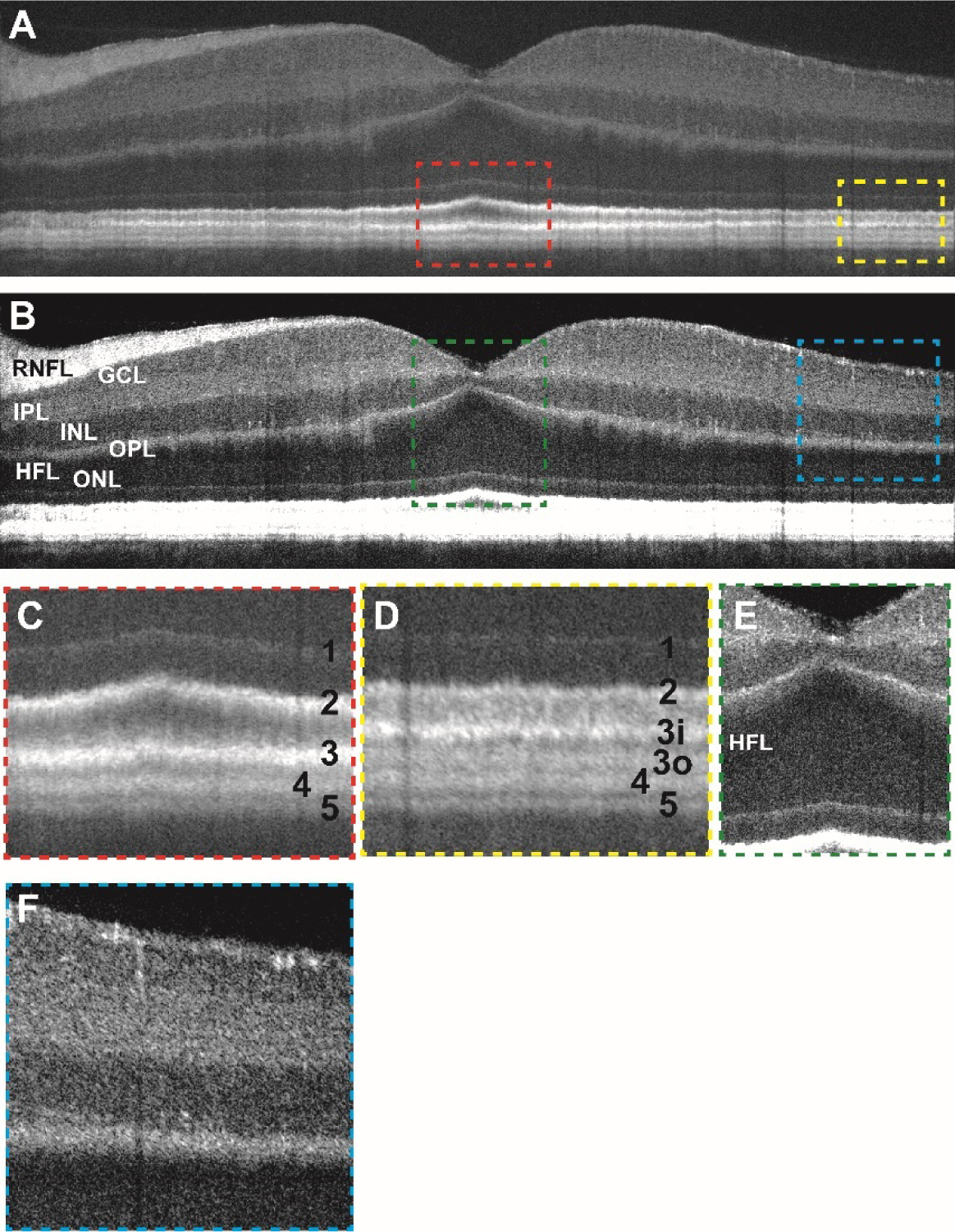
**(a)** Contrast-adjusted VIS-OCT image of a 36-40 age range Caucasian male subject with no known ocular history or retinal pathology highlighting outer retinal features. **(b)** Contrast-adjusted image highlighting inner retinal features. **(c)** Magnified view of foveal outer retinal features seen in panel (a) demonstrating outer retinal banding pattern with outer retinal bands labeled. **(d)** Magnified view of parafoveal outer retina. **(e)** Magnified view of foveal inner retinal features seen in panel (c) (see abbreviations below). **(f)** Magnified view of parafoveal inner retinal features. RNFL: retinal nerve fiber layer, GCL: ganglion cell layer, IPL: inner plexiform layer, INL: inner nuclear layer, OPL: outer plexiform layer, ONL: outer nuclear layer, HFL: Henle’s fiber layer

**Table 1:**
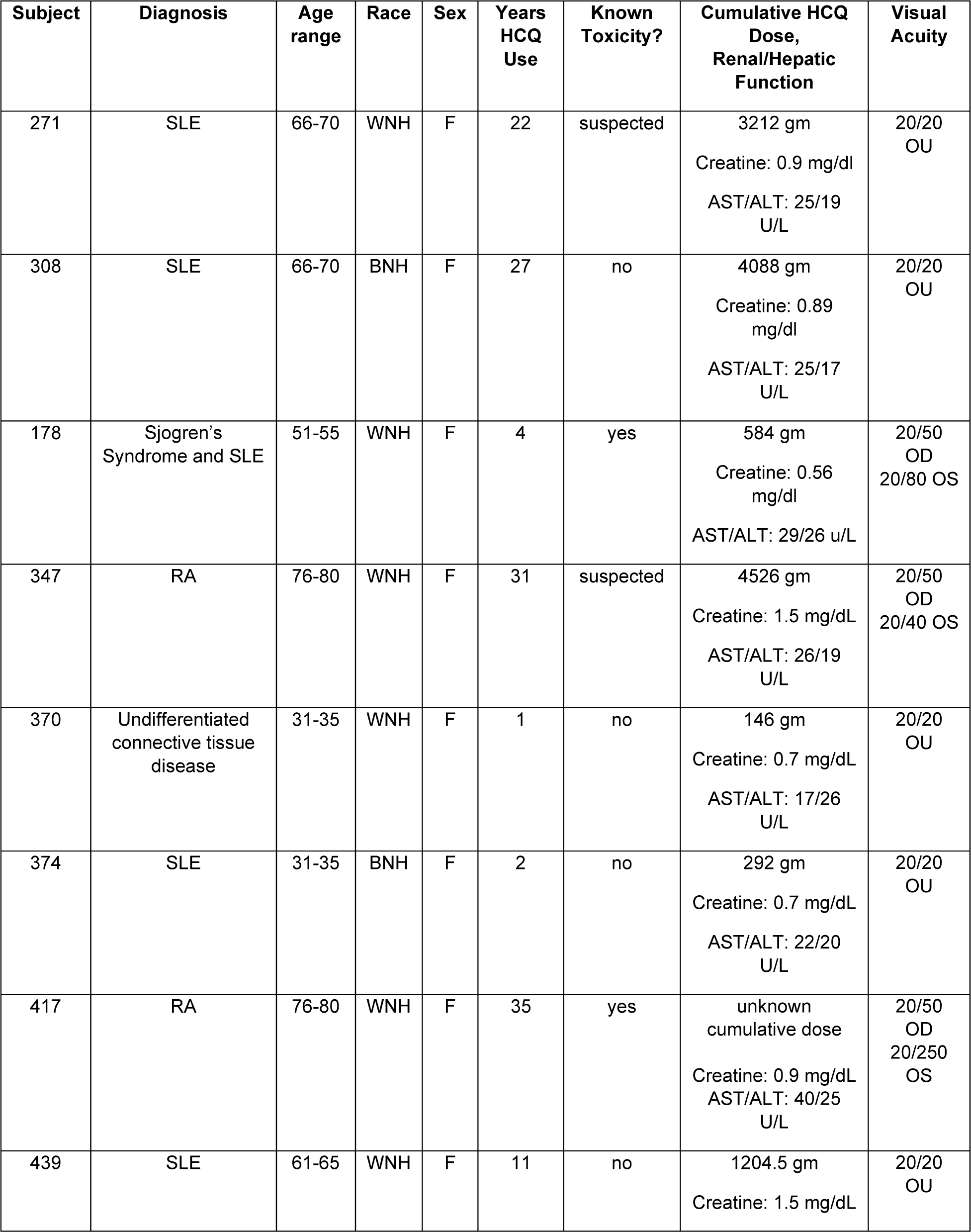
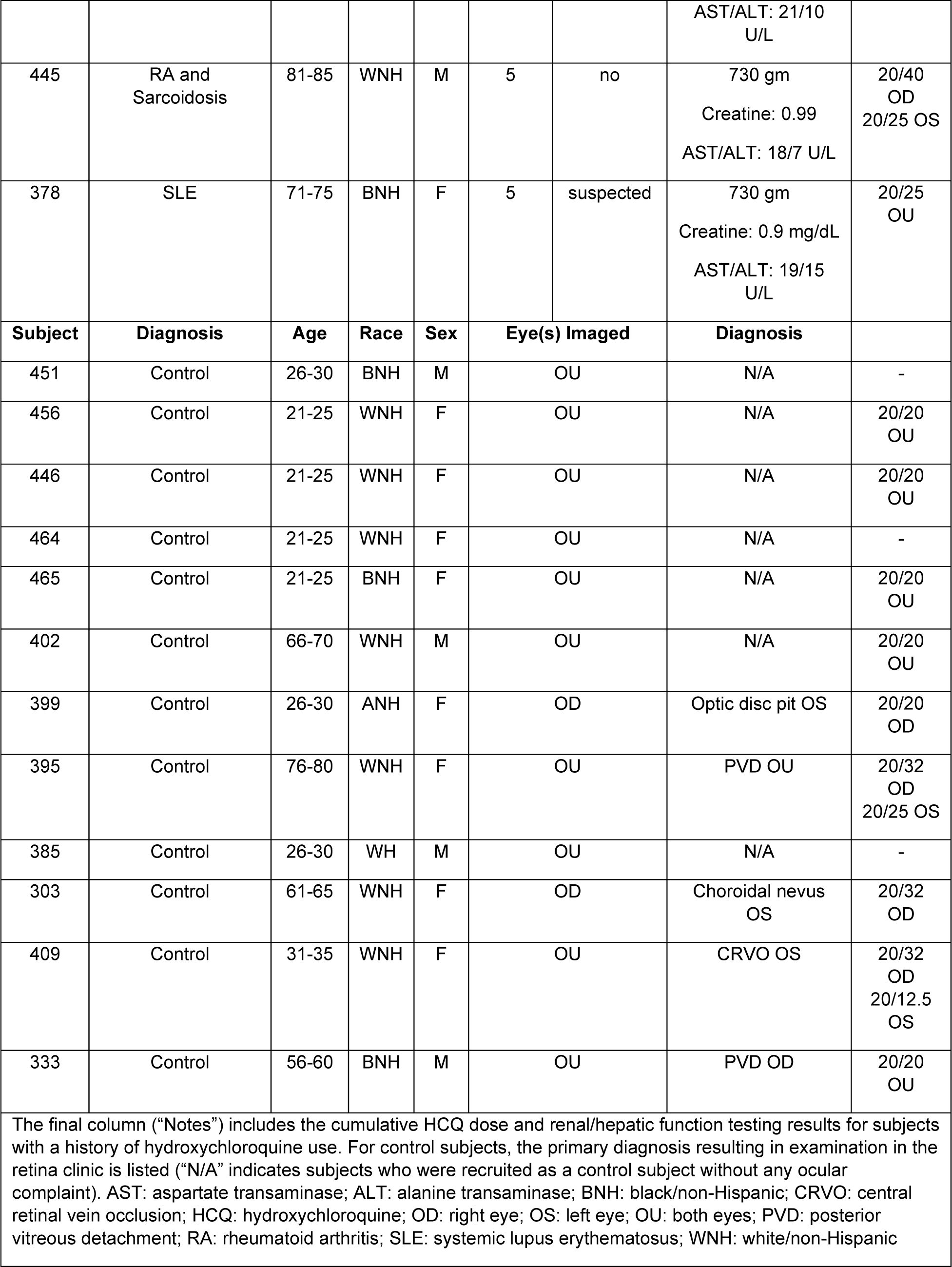
Demographic information of study subjects.

### Imaging Device

The dual-channel VIS-OCT features a visible-light bandwidth ranging from 500 to 650 nm and a near-infrared light bandwidth spanning from 750 nm to 900 nm, with power levels of 0.25 mW and 0.8 mW on the cornea, respectively. Subjects were instructed to fixate using one of two LED displays that served as an external fixation target. A tunable lens was utilized to correct spherical errors. For each eye, the image was initially aligned and optimized under the NIR channel and then promptly imaged with the visible light channel.

### Imaging Protocol

All subjects had standard SD-OCT imaging performed as part of their standard of care assessment. In addition, subjects were scanned using two research OCT devices including a custom-built ultrahigh resolution visible light OCT (VIS-OCT) system and a commercially available SS-OCT system (PlexElite, Carl Zeiss Meditec, Dublin, CA. Imaging on the SS-OCT was performed using a standard 6mm x 6mm raster scan pattern centered on the fovea. The VIS-OCT imaging was performed using a high-definition 4-line radial scanning pattern (32×2048 A-lines x 4 B-scans) as well as multiple high-definition single line scans (64×1024 A-lines x 1 B-scan) in the region of the raster scan pattern from the SS- and SD-OCT devices. For all VIS-OCT scans, 32 or 64 modulated A-lines over ∼0.1 mm orthogonal to the B-scan direction were averaged to produce high-quality images with reduced speckle noise^21,22^. The line rate was 100kHz with a total acquisition time of 2.62 seconds per scan pattern. Images were flattened using a custom algorithm. VIS-OCT and SS-OCT foveal scans were manually registered by using the depth of the foveal center.

### Image Processing for VIS-OCT

We employed a per-A-line noise cancellation algorithm to eliminate baseline light source spectrum, reduce noise, and enhance the signal-to-noise ratio. Dispersion compensation and fast Fourier transform (FFT) were then applied to generate B-scans. To establish the display range, we defined the intensity of background after FFT as the lower boundary and selected the intensity value at 0.05% from the sorted values (from high to low) as the upper boundary. Utilizing these upper and lower boundaries, we applied a logarithmic scale and normalization for image display. Given the substantial dynamic range (exceeding 60 dB for control subjects) between the inner and outer retina, achieving a balanced visual effect proved challenging. Therefore, we increased the image brightness by 40% to enhance the visibility of the inner retina, although this resulted in saturation of the outer retina. Additionally, we manually flattened the B-scans based on band 5 (Bruch’s membrane) **(Supplemental Figure 1**). The intensity of each outer retinal layer was quantified by manually segmenting the boundaries of each layer and calculating the cumulative intensity between the boundaries. Manual segmentation was performed independently by two separate graders and differences were adjudicated by a third grader.

## Results

As described in Table 1, a total of 20 eyes from 12 control subjects (without known retinal pathology) were recruited into this study. VIS-OCT imaging of these subjects demonstrates outer retinal anatomy in finer detail than previously possible with conventional OCT technology. **Figure 1 and Supplemental Figure 2** demonstrate representative VIS-OCT images from control subjects. Within the foveola, five hyper-reflective outer retinal bands are visualized, labeled as bands 1-5 (**Figure 1C**). These bands are putatively identified as the external limiting membrane (ELM, band 1), ellipsoid zone (EZ, band 2), cone photoreceptor outer segment tips (COST, band 3), retinal pigment epithelium (RPE, band 4) and Bruch’s Membrane (BM, band 5). This banding pattern was observed in all control subjects irrespective of patient age. Henle’s fiber layer was visible as a slightly darker region above the outer nuclear layer (**Figure 1B, 1E**). Magnified views of the inner plexiform layer demonstrate sublayers in the perifoveal region (**Figure 1B and 1F**), consistent with previous findings^23^.

In the parafoveal region, outer retinal band 3 consistently divides into two distinct hyperreflective bands (labelled bands 3 inner (3i) and 3 outer (3o)) for a total of six hyperreflective bands (**Figure 1A and 1D**). In accordance with prior anatomical studies and previously published VIS-OCT imaging data^13,21^, these bands are putatively identified as the cone and rod photoreceptor outer segment tips (COST/ROST), respectively. To better characterize these bands, each one was manually segmented, and the mean intensity of each band averaged across three control subjects was plotted as a function of the distance from the fovea (**Figure 2**). Notably, the intensity of band 3i peaks in the foveal region and sharply declines with increasing retinal eccentricity. On the other hand, band 3o is not visible in the foveal region and gradually appears with increasing retinal eccentricity. This pattern mirrors previously published histological studies of cone and rod density^24,25^, supporting the hypothesis that bands 3i and 3o represent cone and rod photoreceptor outer segments tips, respectively. This inverse relationship between bands 3i and 3o was consistent between two graders who independently segmented the layers (**Supplemental Figure 3**).

**Figure 2:**
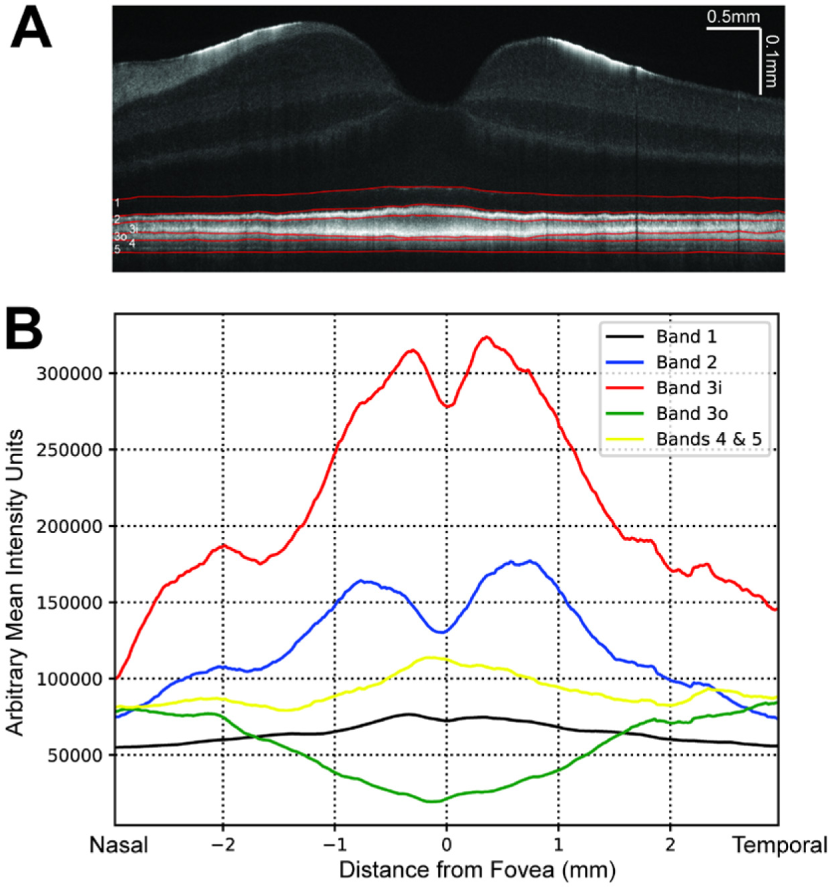
**(a)** VIS-OCT image of control subject demonstrating manual segmentation of outer retinal layers (red). **(b)** Mean intensity of each outer retinal band (summation of intensity between manually segmented outer retinal bands) averaged across three control subjects shown relative to distance from the foveal pit.

A total of 20 eyes from 10 subjects with a history of hydroxychloroquine use were recruited (**Table 1**). These subjects demonstrated a range in severity of hydroxychloroquine toxicity findings (including subjects with suspected toxicity or no evidence of toxicity on prior testing) and were imaged using VIS-OCT. These subjects range in age (33 to 84 years) and length of hydroxychloroquine use (1 to 35 years). The approximate cumulative lifetime hydroxychloroquine dose was calculated, and each patient’s most recent renal and hepatic function testing was recorded if available from their medical records.

**Figure 3** illustrates data from a 61-65 age range female subject with an 11-year history of hydroxychloroquine use and qualitatively unremarkable SD-OCT and SS-OCT scans (**Figure 3B**). The patient had excellent visual acuity (20/20 in both eyes) and denied any subjective vision changes at the time of examination. Hydroxychloroquine toxicity was suspected based on serial SD-OCT mean central subfield thickness measurements showing decreasing thickness in the nasal and temporal ETDRS subfield sectors as described in Melles et al., 2022 (**Figure 3B, 3F**). VIS-OCT images (**Figure 3A, 3C-D**) were compared to a control subject (**Figure 3E**) and demonstrated blunting of most band boundaries as well as essentially complete loss of Band 3i in the parafoveal region. **Figure 3G** illustrates the average intensity profile of the outer retina in the nasal parafovea from 3mm to 6mm eccentricity, corresponding to the outer parafoveal ETDRS subfield (indicated with vertical white dashed lines in **Figure 3A**). There is a marked decrease in the pixel intensity of regions corresponding to band 3i and 3o (red line) relative to the average of three control subjects (black line) as well as in comparison to the intensity of bands 4 and 5, suggestive of damage to the photoreceptor bands. This decrease in band reflectivity (∼5x decrease) is much larger proportionately than the qualitative decrease in the thickness from ELM to Bruch’s membrane illustrated in **Figure 3G**. Also, the mean retinal thickness from the same 3mm to 6mm nasal parafoveal ETDRS subfield is within the normal range relative to a normative database (279 µm in **Figure 3F**).

**Figure 3:**
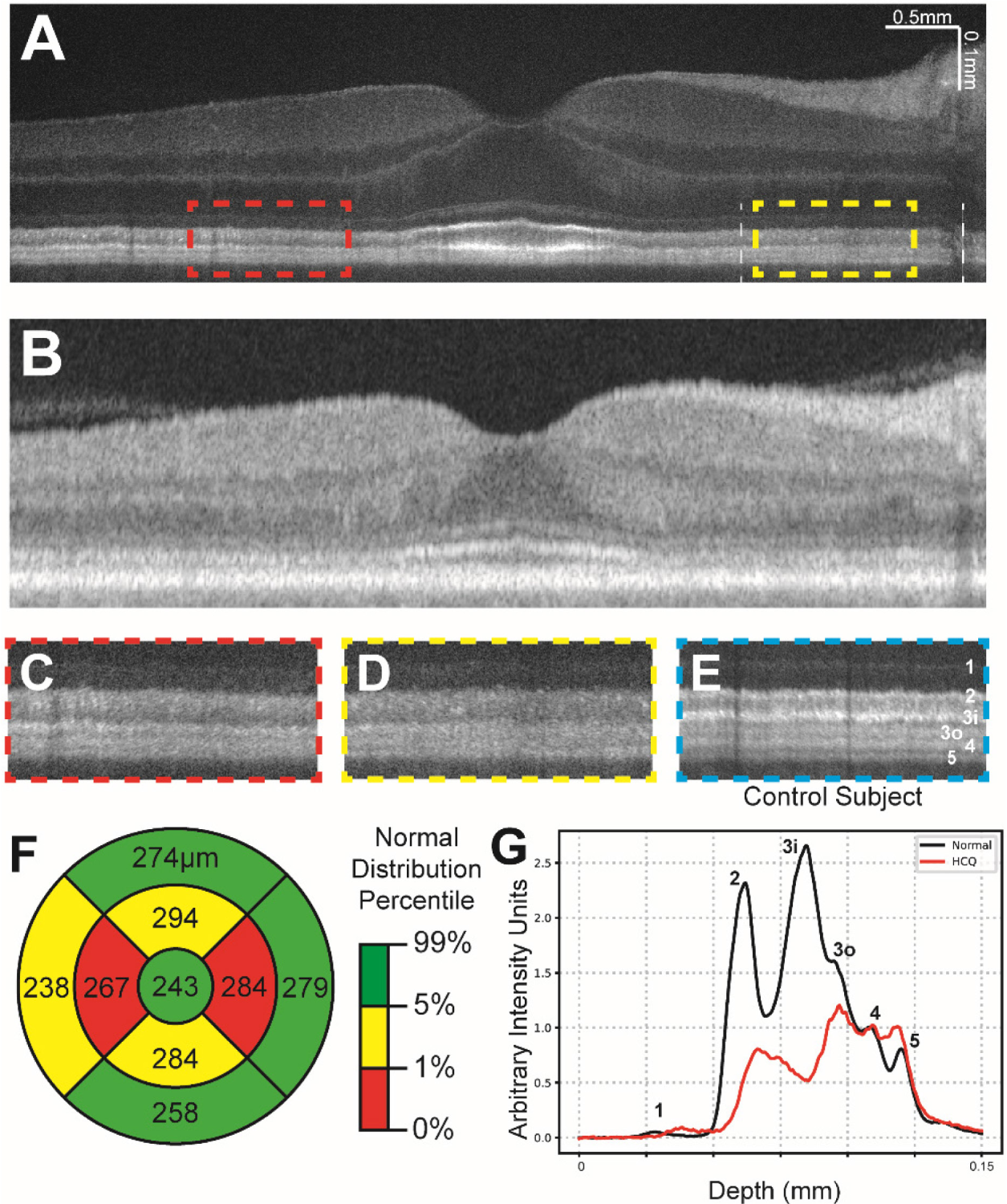
**(a)** Contrast-adjusted VIS-OCT image of a 61-65 age range subject with a history of systemic lupus erythematosus and 11 years of hydroxychloroquine use. **(b)** Swept-source OCT image of same subject in (a). **(c-d)** Magnified view of parafoveal outer retinal features (dashed lines in (a)) demonstrating broadening of band 2 and loss of layer 3i. **(e)** Magnified view of outer retinal features from control subject at similar distance to fovea as c-d, scaled to align with panels c-d. **(f)** Central subfield thickness map of total retinal thickness obtained from swept-source OCT, measurements listed in micrometers. Coloring indicates normal distribution percentile per device manufacturer. **(g)** Averaged A-line signal over nasal 3mm-6mm eccentricity from Panel A between two white lines. The A-line trace signal is extracted from the same region in normal eyes in Figure 1. Both A-lines have zero means and were normalized to RPE band.

**Figure 4** illustrates data from a 71-75 age range female subject with a 5-year history of hydroxychloroquine use and excellent best corrected visual acuity of 20/25 in the right eye and 20/20 in the left eye. She denied any subjective vision changes at the time of the examination. SD and SS-OCT images suggested a very mild parafoveal attenuation of the external limiting membrane and/or ellipsoid zone. Serial SD-OCT central subfield thickness assessments suggested thinning in the nasal and temporal parafoveal ETDRS subfields as described in Melles et al., 2022 (**Figure 4F**). These findings were concerning for early hydroxychloroquine toxicity and the patient was closely monitored while encouraging minimizing the dose of hydroxychloroquine. VIS-OCT imaging demonstrates diffuse attenuation of band 3i and patchy parafoveal attenuation of band 3o, which were not clearly apparent on individual b-scans scans in commercially available devices (**Figure 4A versus 4B**) nor in control subjects (**Figure 4C,D** compared to Figure 4E). Similar to the subject demonstrated in **Figure 3**, there was a marked decrease in the intensity of the photoreceptor bands (particularly bands 3i and 3o) relative to the intensity of bands 4 and 5 (**Figure 4G**). This decrease in band reflectivity was present despite of any qualitative decrease in the thickness from ELM to Bruch’s membrane, as illustrated in Figure 4G. The subject ultimately stopped HCQ due to the significant concern for early toxicity.

**Figure 4:**
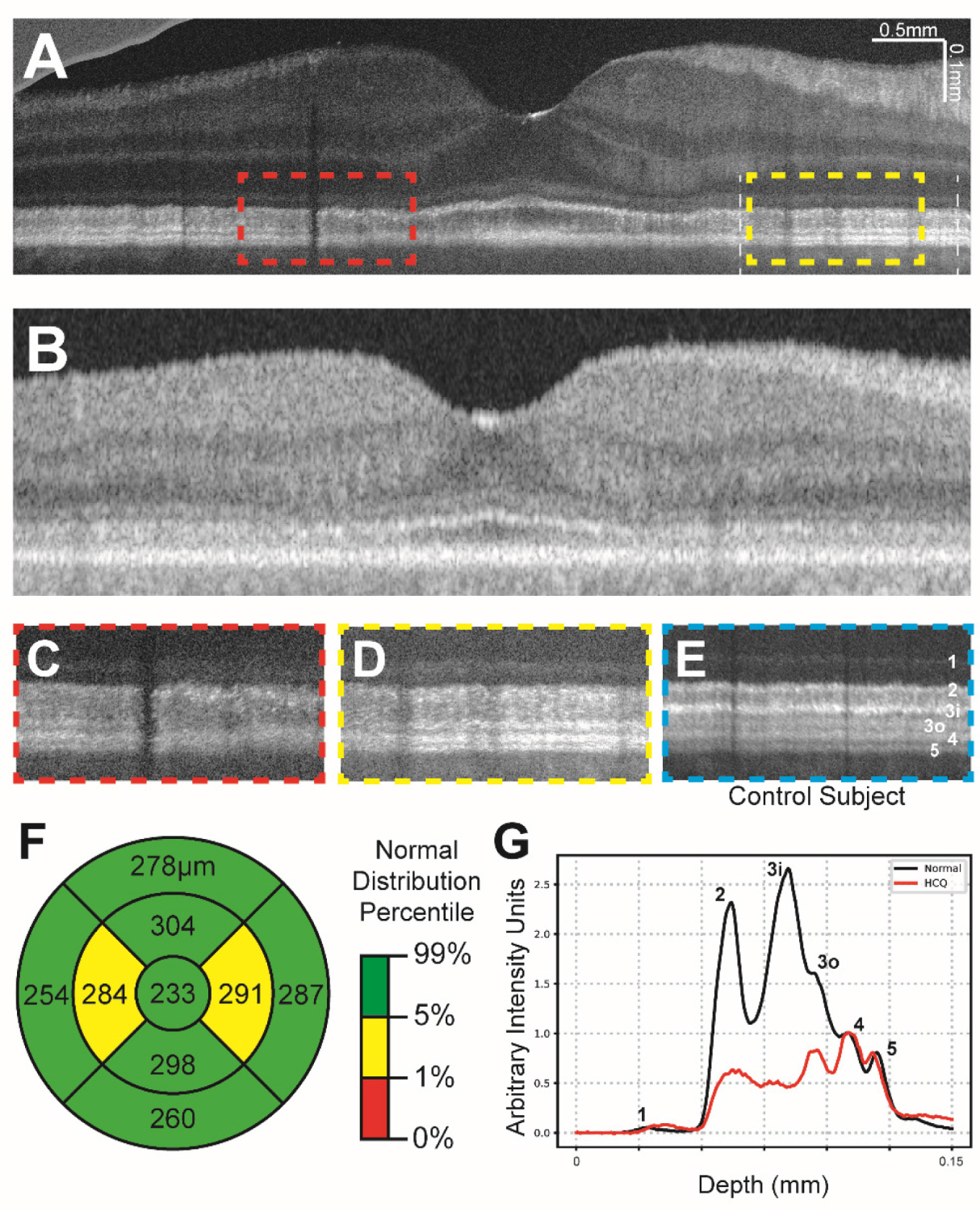
**(a)** Contrast-adjusted VIS-OCT image of 71-75 age range patient with a 5-year history of HCQ use and suspected HCQ retinal toxicity. **(c-d)** Magnified view of parafoveal outer retinal features (dashed lines in (a)) demonstrating broadening of band 2 and loss of band 3i and patchy attenuation of band 3o. **(e)** Magnified view of outer retinal features from control subject at similar distance to fovea as c-d, scaled to align with panels c-d. **(f)** Central subfield thickness map of total retinal thickness obtained from swept-source OCT, measurements listed in micrometers. **(g)** Averaged A-line signal over nasal 3mm-6mm eccentricity from Panel A between two white lines. The A-line trace signal is extracted from the same region in normal eyes in Figure 1. Both A-lines have zero means and were normalized to RPE band.

VIS-OCT imaging of subjects with symptomatic and severe HCQ retinal toxicity demonstrated marked parafoveal attenuation of bands 2, 3i, 3o, and 4 (**Figure 5A)**. Given the severity of damage, this attenuation was also evident with SS-OCT (**Figure 5B**) and with a marked decrease in SD-OCT central subfield retinal thickness as well (**Figure 5F**). At the time of imaging, the patient’s best-corrected visual acuity was 20/63 in the right eye and 20/80 in the left eye and the patient had been previously diagnosed with severe HCQ toxicity. The medication had been discontinued prior to our examination. Notably, even in this subject, there was more severe attenuation of band 3i compared to any other band with increasing eccentricity beyond the parafovea where clear atrophy was not evident. While all bands were less distinct than normal, bands 1, 2 and 3o were particularly diffuse when compared with a control subject (**Figure 5C, D** compared for E). In no case was the attenuation of any band more severe than the attenuation of Band 3i (**Figure 5G**). As in the previous case, this decrease in band reflectivity was present despite of any qualitative decrease in the thickness from ELM to Bruch’s membrane, as illustrated in **Figure 5G**.

**Figure 5:**
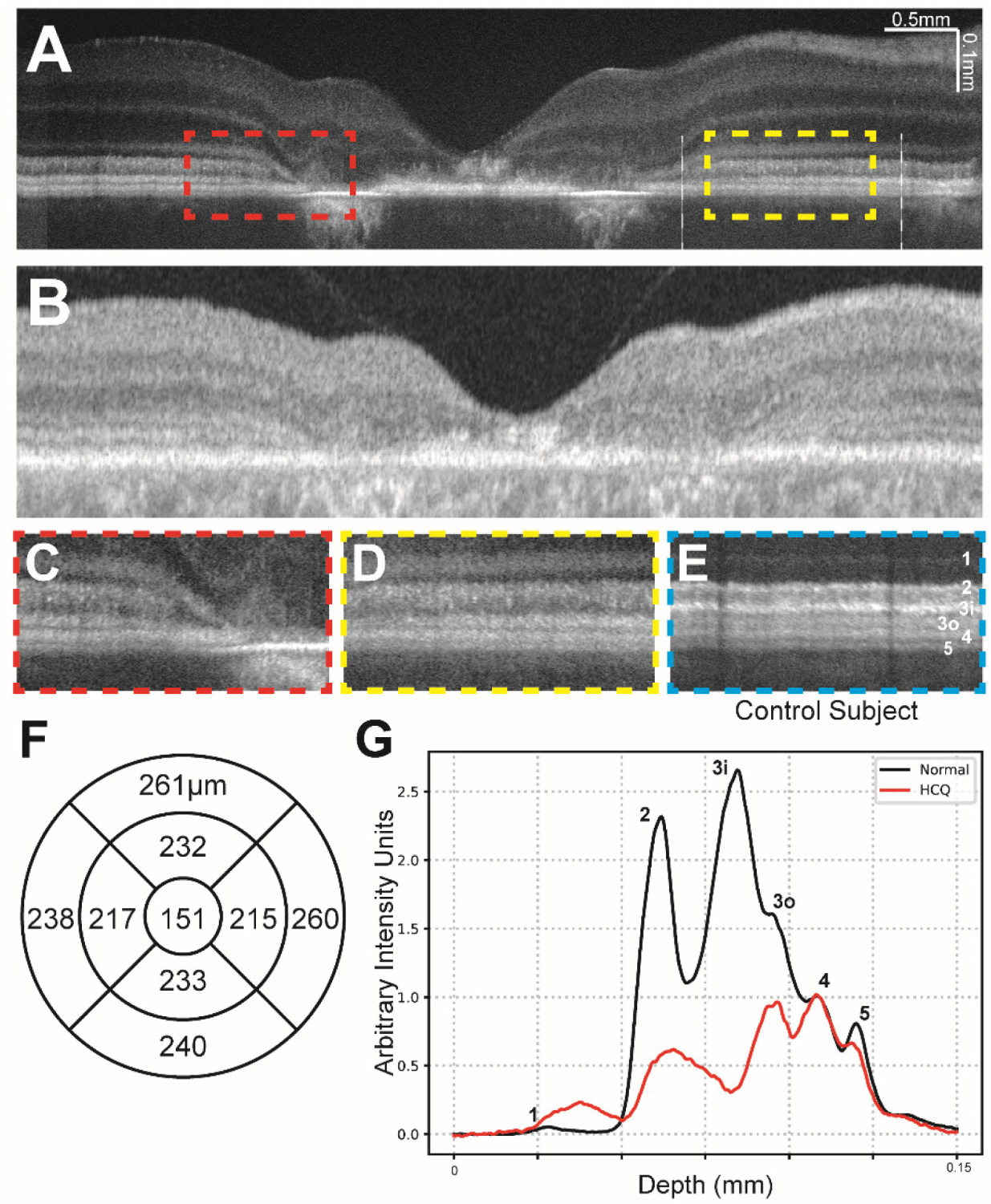
**(a)** Contrast-adjusted VIS-OCT image of 51-55 age range patient with a 4-year history of HCQ use with severe HCQ retinal toxicity and no other known retinal pathology. **(c-d)** Magnified view of parafoveal outer retinal features (dashed lines in (a)) demonstrating diffuse loss of bands 2, 3i, 3o, and 4. **(e)** Magnified view of outer retinal features from control subject at similar distance to fovea as c-d, scaled to align with panels c-d. **(f)** Central subfield thickness map of total retinal thickness obtained from swept-source OCT, measurements listed in micrometers. Normative data not available. **(g)** Averaged A-line signal over nasal 3mm-6mm eccentricity from Panel A between two white lines. The A-line trace signal is extracted from the same region in normal eyes in Figure 1. Both A-lines have zero means and were normalized to RPE band.

## Discussion

Using ultrahigh resolution visible light OCT, we demonstrate outer retinal banding patterns that are not clearly or reliably visible with commercially available SD or SS-OCT. Most importantly, these changes in banding pattern reflectivity are much larger in magnitude than changes in retinal thickness measured qualitatively or quantitatively on SD-OCT or SS-OCT. We also demonstrate that the outer retinal band intensity profiles on VIS-OCT in healthy controls are similar to the known density profiles of rods and cones from histologic studies. In control subjects, we consistently identify five outer retinal bands in the foveola (Bands 1, 2, 3, 4, and 5) and six bands in the parafovea (Bands 1, 2, 3i, 3o, 4, and 5). These bands putatively represent (1) the external limiting membrane, (2) the ellipsoid zone, (3i) the cone outer segment tips, (3o) the rod outer segment tips, (4) the retinal pigment epithelium and (5) Bruch’s membrane. These data strongly suggest that VIS-OCT imaging can distinguish rod- and cone-specific retinal anatomy non-invasively.

We demonstrate the utility of ultrahigh resolution VIS-OCT to detect sub-clinical changes in the outer retinal band reflectivity corresponding to photoreceptor outer segments in asymptomatic subjects at high risk of hydroxychloroquine toxicity. Specifically, we observe that Band 3i (corresponding to the putative cone outer segment tips) is consistently and most severely attenuated in subjects at high risk of toxicity and in whom serial SD-OCT measurements from commercial devices demonstrate retinal thinning. Notably, this latter finding of serial thinning has been implicated in HCQ toxicity^6^. We hypothesize that attenuation of Band 3i is the earliest sign of HCQ toxicity and may be readily detectable on a single visit VIS-OCT while serial SD-OCT measurements over months or years are needed to detect decreasing thickness trends. In our individual VIS-OCT scans, the attenuation of VIS-OCT banding reflectivity is present despite normal retinal thickness as measured with central subfield thickness using SD-OCT when compared with age-matched control subjects. We hypothesize that this outer retinal attenuation of band 3i is the earliest known marker of hydroxychloroquine toxicity. Our analyses assessing the cumulative intensity of bands 3i and 3o at increasing retinal eccentricities in **Figure 2** closely align with previously published anatomical studies quantifying cone and rod densities^24^, supporting the hypothesis that these bands represent the cone and rod photoreceptor outer segments, respectively.

The mechanism of hydroxychloroquine retinopathy is not clearly understood, though prior studies have suggested multiple potential mechanisms^26^. A study by Xu et al. demonstrates that both chloroquine and hydroxychloroquine inhibit organic anion transporting peptide 1A2 (OATP1A2), which mediates uptake of all-*trans*-retinoic acid in the retinal pigment epithelium in the visual cycle.^27^ This may lead to toxicity of both photoreceptors as well as the retinal pigment epithelium. Animal studies have revealed the binding of chloroquine to pigmented retinal structures, including the retinal pigment epithelium^28^. It is unclear why band 3i appears to be attenuated first in subjects on HCQ followed by band 3o, though this finding suggests a tendency for cone photoreceptors to be preferentially over rods. In more severe stages, as shown in **Figure 5**, there appears to be attenuation of the remaining outer retinal bands including the retinal pigment epithelium, favoring a mechanism of toxicity affecting both the RPE as well as the photoreceptors.

Recent work has measured ellipsoid zone attenuation with SD-OCT to detect and quantify HCQ toxicity^29,30^. Our results demonstrate the ability of VIS-OCT imaging to detect early HCQ toxicity with attenuation of the photoreceptor bands, which are not easily visible with SD-OCT or SS-OCT, prior to EZ attenuation. We demonstrate that early HCQ toxicity is characterized mainly by attenuation of bands 3i and 3o, with more severe toxicity affecting the EZ and the retinal pigment epithelium. This corroborates earlier studies that suggest damage occurs to the photoreceptors^4,28^. As demonstrated in Figures 3 and 4, early toxicity is often not apparent on individual line scans with SS-OCT (or SD-OCT), requiring averaging of retinal thickness across a large region to observe retinal thinning on serial thickness maps. Using VIS-OCT, these changes are apparent on foveal line scans, and may assist with earlier diagnosis of HCQ toxicity.

It is important to note that there are limitations to VIS-OCT imaging compared to SS-OCT and conventional FDA approved devices. First, the brightness of the visible light used to acquire VIS-OCT images can be distracting to patients, particularly when utilizing lengthy imaging protocols or in patients who are light sensitive. Additionally, while its increased resolution allows for visualization of Bruch’s membrane, visible light is limited in its ability to penetrate beyond Bruch’s membrane and visualize structures within the choroid, which may limit its utility in the diagnosis of choroidal pathologies.

Due to our small sample size and our study being limited to subjects with HCQ use and toxicity, it remains unclear whether the pattern of changes described can also be seen in other retinal diseases or whether they are specific to HCQ toxicity. Our findings support previous work suggesting that anatomically detectable damage to photoreceptors precedes similar damage to the RPE^1,4^. Prospective studies and analyses with larger sample sizes will be necessary to further characterize these changes. Additionally, as previously described, the pattern of retinal toxicity and vision loss associated with HCQ use varies in patients of Asian descent, and additional work is necessary to determine whether VIS-OCT can demonstrate this difference or provide further insight into its pathogenesis. Prior work has also demonstrated that in a minority of patients, early HCQ toxicity is detected on visual field testing prior to clear SD-OCT changes although this is likely due to the limited resolution of single SD-OCT scans to reliably demonstrate changes in the outer retinal layers as we have shown above.^31^ Further work with a larger sample size is necessary to determine whether changes on VIS-OCT can reliably be detected prior to visual field changes.

## Data Availability

All data produced in the present study are available upon reasonable request to the authors

## Abbreviations

OCT: optical coherence tomography
VIS: visible light
SS: swept source
SD: spectral domain
HCQ: hydroxychloroquine
ELM: external limiting membrane
EZ: ellipsoid zone
COST: cone outer segment tips
ROST: rod outer segment tips
RPE: retinal pigment epithelium
BM: Bruch’s membrane

---

**Supplemental Figure 1:**
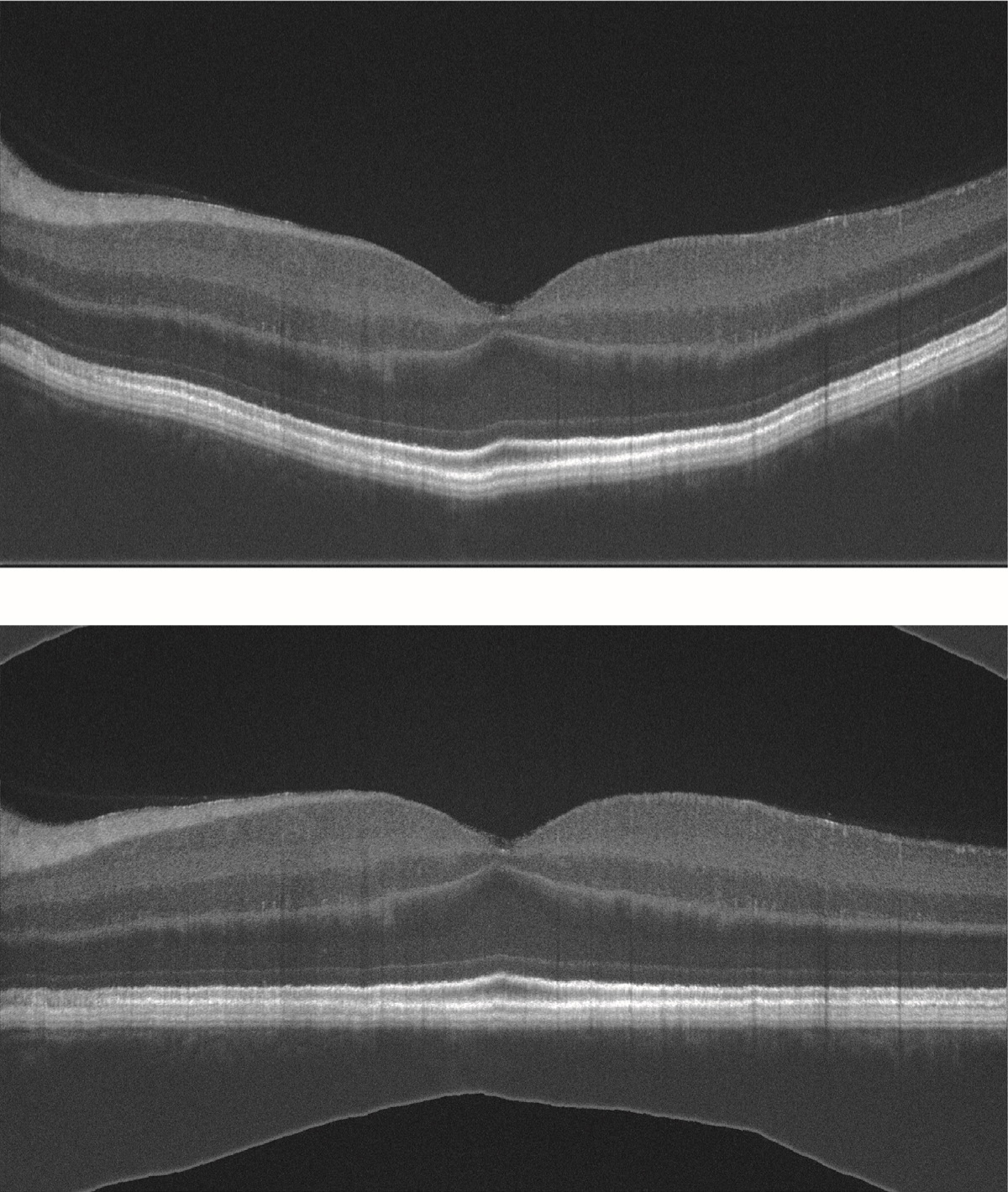
**(a)** Sample raw output of foveal line scan from VIS-OCT device prior to image processing. **(b)** Foveal line scan following image flattening and contrast adjustment as described in Methods.

**Supplemental Figure 2:**
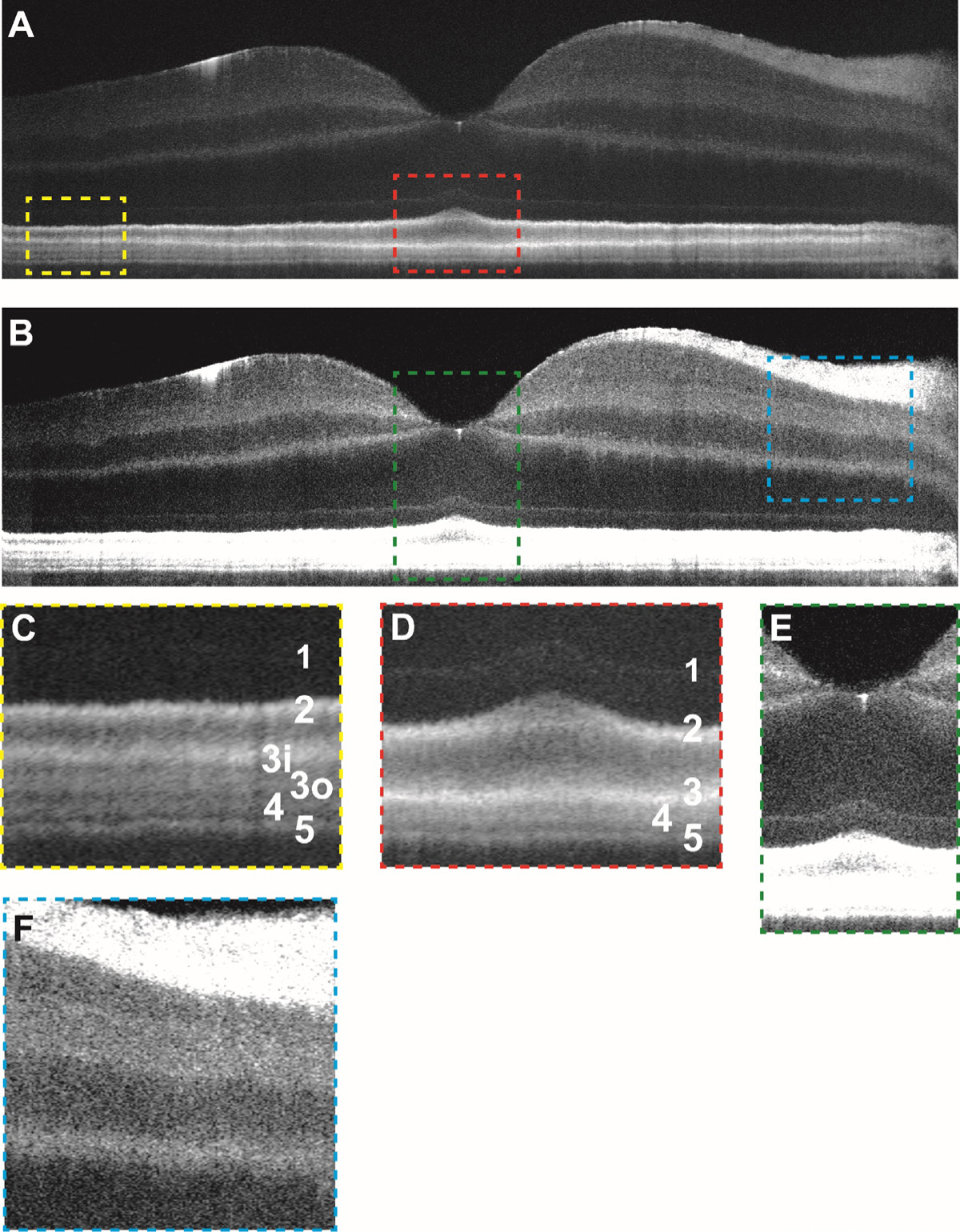
**(a)** Contrast-adjusted VIS-OCT image of a 26-30 age range Caucasian male subject with no known ocular history or retinal pathology highlighting outer retinal features. **(b)** Contrast-adjusted image highlighting inner retinal features. **(c)** Magnified view of foveal outer retinal features seen in panel (a) demonstrating outer retinal banding pattern with outer retinal bands labeled. **(d)** Magnified view of parafoveal outer retina. **(e)** Magnified view of foveal inner retinal features seen in panel (c) (see abbreviations below). **(f)** Magnified view of parafoveal inner retinal features.

**Supplemental Figure 3:**
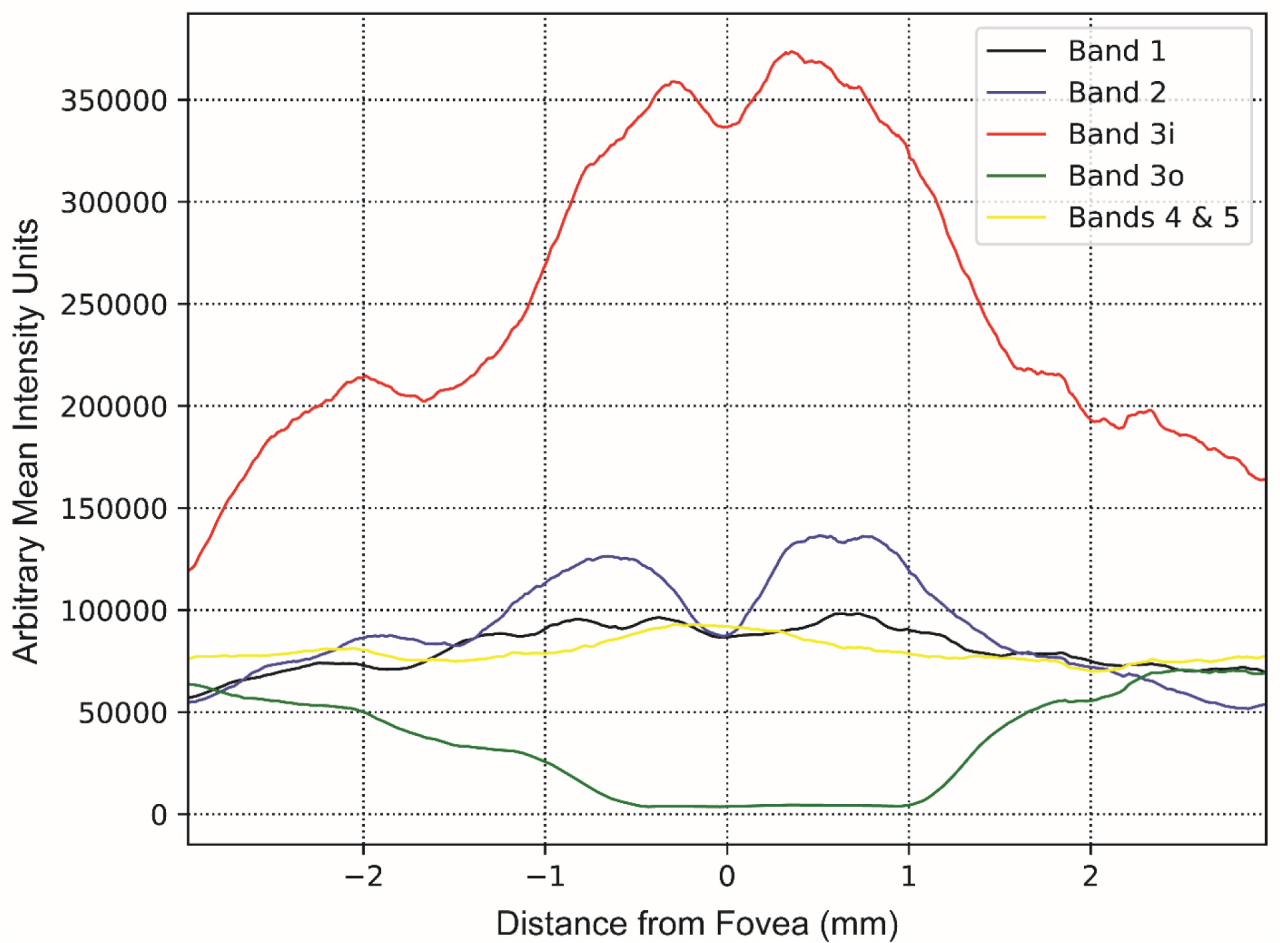
Mean intensity of each outer retinal band (summation of intensity between manually segmented outer retinal bands) averaged across three control subjects shown relative to distance from the foveal pit for VIS-OCT scan displayed in Figure 2A. Manual segmentation of outer retinal bands performed independently by second grader.

